# Is climate a curse or a bless in the Covid-19 virus fighting?

**DOI:** 10.1101/2020.09.04.20182998

**Authors:** Olivier Damette, Clement Mathonnat, Stephane Goutte

## Abstract

Faced with the global pandemic of Covid-19, we need to better understand the links between meteorological factors and the virus and investigate the existence of potential seasonal patterns. In the vein of a recent empirical literature, we reassess the impact of weather factors on Covid-19 daily cases in a panel of advanced and emerging countries between January the first and 28th May 2020. We consider 5 different meteorological factors and go further previous studies. In addition, we give a short-run and medium/long-run time perspective of the dramatic outcomes of the pandemic by both considering infected people (short-run) and fatalities (long-run). Our results reveal that the choice of delays and time perspective of the effects of climatic factors on the virus are crucial as well as Covid-19 outcomes can explain the discrepancies in the previous literature. For the first time, we use a dynamic panel model and consider two different kinds of channels between climate and Covid-19 virus: 1) direct/physical factors related to the survivals and durability dynamics of the virus in surfaces and outdoors and 2) an indirect factor through human behaviors and individual mobility – walking or driving outdoors – to capture the impact of climate on social distancing and thus on Covid-19 outcomes. Our model is estimated *via* two different estimators and persistence, delays in patterns, nonlinearities and numerous specifications changes are taken into account with many robustness checks. Our work highlights that temperatures and, more interestingly, solar radiation – that has been clearly undervalued in previous studies – are significant climatic drivers on Covid-19 outbreak. Indirect effects through human behaviors i.e interrelationships between climatic variables and people mobility are significantly positive and should be considered to correctly assess the effects of climatic factors. Since climate is *per se* purely exogenous, climate tend to strengthen the effect of mobility on virus spread. The net effect from climate on Covid-19 outbreak will thus result from the direct negative effect of climatic variables and from the indirect effect due to the interaction between mobility and them. Direct negative effects from climatic factors on Covid-19 outcomes – when they are significant – are partly compensated by positive indirect effects through human mobility. Suitable control policies should be implemented to control the mobility and social distancing.

## Introduction

Faced with the global pandemic of a new coronavirus, we need to better understand the main factors that may influence the Covid-19 virus spread. In the Northern Hemisphere, it has been widely hoped that the transition to spring or summer associated with higher temperatures could slow or stop the spread of the pandemic. This sentiment has been based on the principle that the flu or influenza incidence is increasing during December to February periods when temperature and humidity levels are likely to be at lower levels but is low during summer times (1). Over the longer term, as more people develop immunity, some researchers suggest that COVID-19 may likely fall into a seasonal pattern similar to those seen with diseases caused by other coronaviruses. However, pandemic viruses can have different behave as underlined for the pandemic influenza, such as 2009 A/H1N1A or the Spanish flu, at least, in the short-run (1). Nonetheless, some hints from recent lab experiments or statistical studies suggest that increased temperature and humidity may reduce the viability of the SARS-CoV-2.

Indeed, several lab experiment studies found evidence that for other human coronaviruses, the duration of persistence is shorter in warm conditions (2) and tend to confirm the existence of seasonal patterns. (3) demonstrate that the stability of HCoV-19 and SARS-CoV-1 under the experimental circumstances tested is similar. (4) have confirmed for the previous SARS coronavirus that the virus viability was rapidly lost at higher temperatures and relative humidity levels, also leading to different epidemic curves in countries with subtropical and tropical areas, also considering air-conditioned environments. Finally, these results are in accordance with other previous lab experiments on different viruses such as gastroenterit virus and mouse heptatit virus (5) to determine the effects of air temperature and humidity on pathogenic viruses such as the SARS-Cov and confirm the role of high temperatures (20C and more importantly at 40C) as well as the existence of non-monotonic relationships. Other studies (6-7) show that cold and dry conditions increase the transmission of the virus.

Recently, a few statistical studies estimate and simulate how seasonal changes in temperature might influence the trajectory of COVID-19 in cities around the world and especially in US. A few robust previous studies (8-10) concluded that climate may be a marginal significant driver in evaluating the course of the Covid-19 epidemics. However, despite several new empirical investigations, the conclusions of the literature about the climate impact is still mixed and real observations do not validate previous projections. Negative effects of high levels of temperatures and humidity found in some recent empirical studies (11-15) seem consistent with studies about the effect of physical factors on the virus and its survival rate conducted in experimental works. Regions with low humidity and average temperatures between 40 and 50 degrees Fahrenheit are likely to have a more important spreading of the virus (10); above 25 degrees, there is a strong association between temperatures and reduced transmission rates with the largest effect is 30 to 40 percent reduction in the rate but in most locations, even 40 percent still leave Covid-19 climbing at an exponential rate and this situation is only seen for very hot and humid conditions (13). (14) show that although cases of COVID-19 are reported all over the world, most outbreaks display a pattern of clustering in relatively cold and dry areas.

However, this positive association between climate conditions and Covid-19 is a controversial debate and the relationship may be weak (16) and even if one assumes that SARS-Cov-2 is as sensitive to climate as other seasonal viruses, summer heat still would not be enough right now to slow down its rapid initial spread through the human population. (17) find evidence of a positive association between daily death counts and diurnal temperature range, (18) a positive linear relationship between Covid-19 cases and mean temperatures but find no clear evidence that the counts of Covid-19 cases are reduced when the weather is warmer. (19) do not find evidence of an association between relatively high temperatures (up to 20C) and the spread rate of the virus. (20) are very doubtful about the existence of significant relationships between high absolute humidity and the survival of this new virus.

Can the lack of controls included in the regressions explain these different empirical results? (21) show that Covid-19 growth rates peaked in temperate regions of the Northern Hemisphere with mean temperature of 5C, and specific humidity of 4-6 g/m3 during the outbreak period by controlling for population size, density and health expenditure during January-March 2020.

Beyond the previous controversial conclusions, we find that there is a lack of work about other climatic factors (solar radiation, wind and precipitations). Precipitations and wind have generally a positive impact on the transmission rate, that could result from people spending more time indoors. (10) consider wind speed (log of Km/hour), precipitations (log of millimeters) and ultraviolet index (25 milliwatts/m2) as well as squared ultraviolet index as potential drivers of the Covid-19 reproduction number and find a U-shaped relationship between UV index and the transmission rate. The UV may help more temperate countries during summer but increase risks in equatorial regions with very high levels of UV exposure. (22) and (23) are in accordance with a negative association between UV levels and Covid-19 cases.

The wind speed is likely to be another determinant of Covid-19 spread since human saliva-disease-carrier droplets may travel up to unexpected considerable distances depending on the wind speed (24). (25) assume that wind speed can affect droplets stability in the environment or the survival of viruses, like air temperature, and as a result, the transmission rate. Although wind speed is not an important factor if modeled as the only explanatory variable, it represents a necessary factor in their final model. For Turkey, (26) find that the 14-day lag of the average wind speed has the highest (positive) correlation with the number of cases. This wind speed effect is also significant for (10) and (24) but not for Oliveiros et al. (27).

## Results

### Preliminaries

Preliminaries (Fig. 1 and Fig. 2) outlining the direct relationship between the mean temperatures and the infected cases (7 lags) and fatality ratios (28 lags) respectively reveal that the relationships contemplated in some previous studies are not self-evident. We also check our results with more longer delays and qualitative results (SI Appendix, Robustness Check) are not very sensitive to the lag choice in this context.

**Figure 1.**
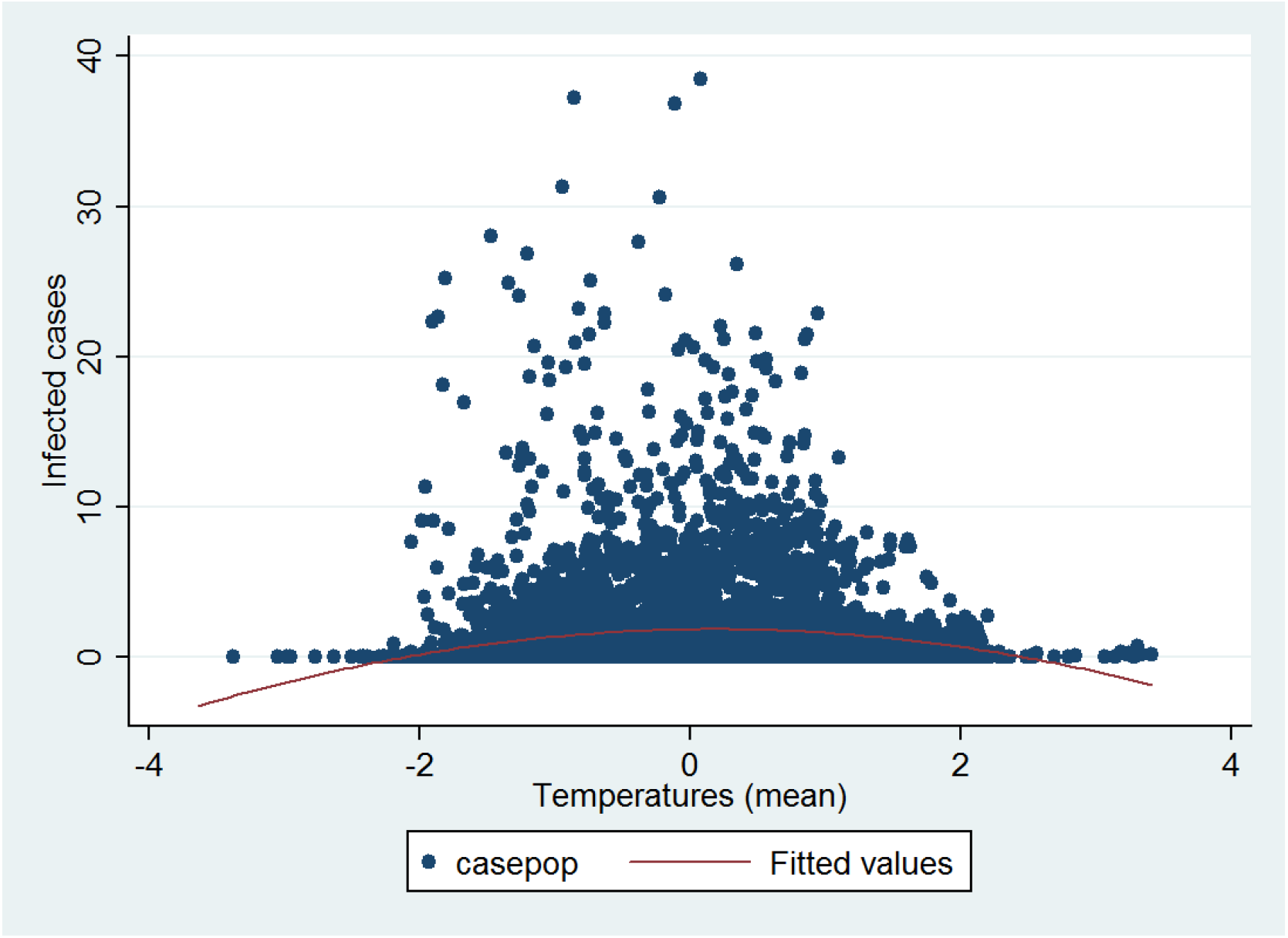
Infected cases and lagged 7 days temperatures (mean)

**Figure 2.**
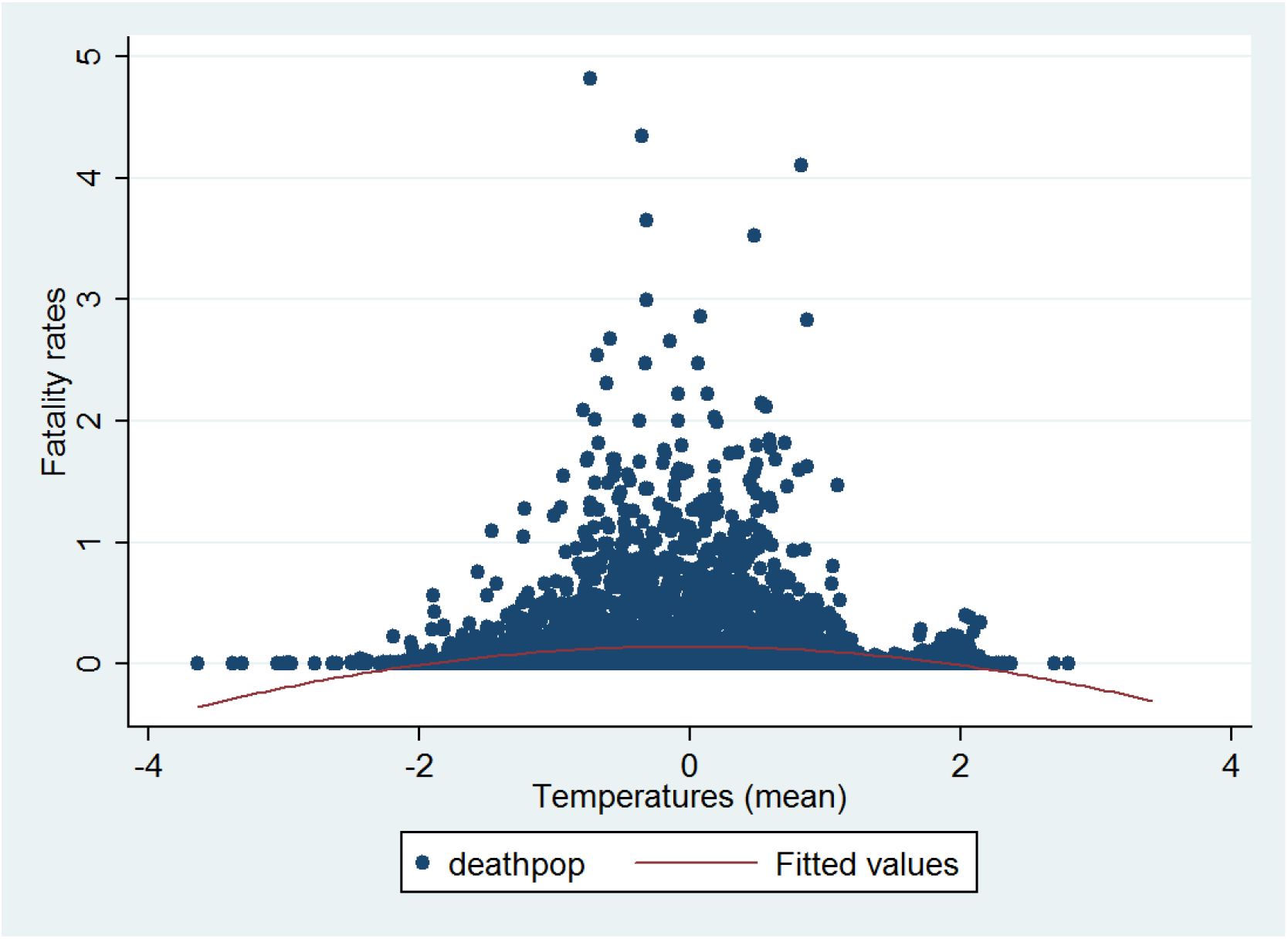
Fatality rate and lagged 28 days temperatures (mean)

In contrast, the relationship between mobility (driving) and temperatures (Fig. 3) outlines a negative -quite nonlinear-relationship between climate and the mobility index. Thus, when the temperature increase, the mobility index (walking as well as) tends to decrease for low levels of temperatures. For higher levels of temperatures, the mobility increases since people go outdoors for leisure activities for example. Using solar radiation instead lead to similar conclusions.

**Figure 3.**
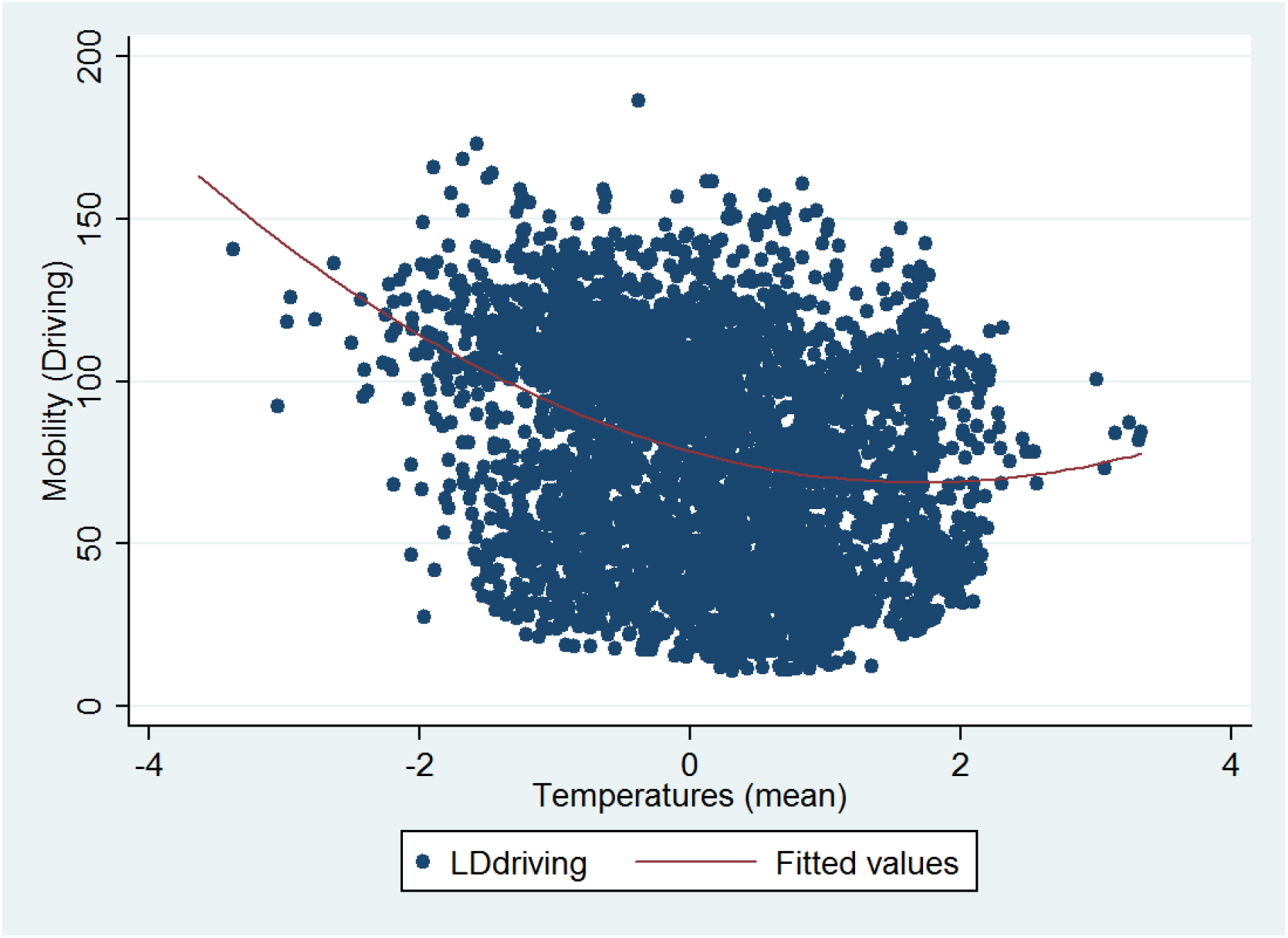
Temperature and mobility (contemporaneous values)

Preliminary analysis tend to suggest that the direct effect of climate on Covid-19 spread is not completely clear: different forms of nonlinearity should be investigated. However, indirect channels through mobility – propensity of people to move outdoors, driving, walking for example – are probably correlated with climatic conditions. It is thus a potential channel to consider when we want to explain the links between climate and the Covid-19 virus.

### Direct climatic factors effects

Our baseline regressions (SI Appendix, Tables S3 and S4) show that temperature and humidity levels – the most studied factor in the previous literature – have a significant negative effect on Covid-19 infected cases ratio. As a consequence, these two factors are likely to reduce the spread of the virus through a direct effect as suggesting by a large part of the previous literature. The same results are derived, albeit to a lesser extent, for precipitations and wind speed. Results are robust to lags choice and to estimator choices. Therefore, these first estimates tend to corroborate the conclusions of a part of the previous statistical literature about a small but significant effect of the climatic factors – mainly temperature and humidity but not only – as reducing drivers of the Covid-19 outbreak.

However, when we turn from infected cases to fatalities taking account a 28 lags delay (Table 1 and SI Appendix, Tables S5-S6), direct climatic effects tend to disappear. Climatic effects are thus not clear-cut when fatalities, and thus long-run perspective, are considered. We also produce some robustness checks by increasing the lags for both cases and fatalities (SI Appendix) from 28 to 42 days. Overall, these results tend to lower the conclusions about the significant effects of climatic variables and show that contrasting conclusions obtained so far in the literature could be explained by the lags chosen to study the effects of climate variables on Covid-19 outcomes. The choice of the different Covid-19 outcomes as endogeneous variables is also not neutral.

**Table 1.** Direct climatic variables effects on Covid-19 fatalities: DFE estimates

| VARIABLES | (1.)<br>deathpop | (2.)<br>deathpop | (3.)<br>deathpop | (4.)<br>deathpop | (5.)<br>deathpop |
| --- | --- | --- | --- | --- | --- |
| deathpop(-1) | 0.629***<br>(0.0757) | 0.629***<br>(0.0760) | 0.629***<br>(0.0755) | 0.629***<br>(0.0755) | 0.628***<br>(0.0760) |
| deathpop(-14) | 0.0931**<br>(0.0401) | 0.0928**<br>(0.0397) | 0.0955**<br>(0.0399) | 0.0940**<br>(0.0397) | 0.0960**<br>(0.0402) |
| casepop(-14) | 0.0104**<br>(0.00443) | 0.0104**<br>(0.00442) | 0.0104**<br>(0.00445) | 0.0105**<br>(0.00444) | 0.0105**<br>(0.00447) |
| temperature(-28) | -0.00463<br>(0.00473) |  |  |  |  |
| humidity(-28) |  | -0.00239<br>(0.00745) |  |  |  |
| radiation(-28) |  |  | -0.00674<br>(0.00467) |  |  |
| precipitation(-28) |  |  |  | 0.00324<br>(0.00311) |  |
| windspeed(-28) |  |  |  |  | 0.00815**<br>(0.00317) |
| Time | 0.000113<br>(9.33e-05) | 7.46e-05<br>(8.65e-05) | 0.000184*<br>(0.000108) | 7.19e-05<br>(7.29e-05) | 9.32e-05<br>(7.91e-05) |
| Constant | 0.0127*<br>(0.00654) | 0.0167***<br>(0.00517) | 0.00550<br>(0.0117) | 0.0169***<br>(0.00578) | 0.0144**<br>(0.00573) |
| Observations | 4,120 | 4,120 | 4,120 | 4,120 | 4,120 |
| R-squared | 0.560 | 0.560 | 0.560 | 0.560 | 0.561 |
| Number of countries | 40 | 40 | 40 | 40 | 40 |

Robust standard errors in parentheses
\*\*\* p<0.01, \*\* p<0.05, \* p<0.1

When all climatic variables are simultaneously added in our econometric model, the results are again mixed and the significant single effects of climatic factors are not clear although temperatures and solar radiation seem to be more robustly correlated with the pandemic (SI Appendix, Table S7).

### Testing for interactions between climatic variables

We next turn to an extended model that consists in adding interacted climatic variables to our benchmark model in order to explicitly test for potential interactions between them: *temperature* * *radiation* and *temperature* * *humidity*. Indeed, these variables are the most strongly correlated between them. Results (SI Appendix, Tables S8-S9) highlight significant interrelationships between the climatic factors. In particular, the relationship between temperature and solar radiation is significant and seems particularly robust in most specifications, whatever the Covid-19 outcome considered. Nevertheless, we notice that the single direct effects of temperatures and solar radiation partially disappear. More importantly, a simultaneous and combinated increase of temperatures and solar radiation is associated with a significant decrease of cases and fatalities. The same result is derived for a combination of high temperatures and humidity but only for infected cases in the MG estimates. Thus, these results rather confirm the expected simultaneous direct effects of temperatures and humidity on Covid-19 cases, as assumed in some lab experiments studies.

### Testing for thresholds effects in climate variables

Our baseline specifications assume that the statistical relationship between meteorological factors and Covid-19 outcomes is linear. We have also investigated the existence of thresholds effects by testing quadratic functions. Indeed, as shown for instance by (10) among others, temperatures (when surpassing 25C for example) or ultraviolet level are likely to exhibit nonlinear patterns. Overall, our results (SI Appendix, Table S10) reveal no threshold evidence except for the solar radiation variable. Concerning the infected cases regressions, ie the short-run effects of weather, temperatures and humidity have a significant and negative effect on the number of infected cases, but their respective thresholds coefficients are insignificant. In addition, albeit the solar radiation variable is not significant, we notice a significant and negative threshold coefficient associated with this variable. This result thus outlines the existence of an inverted U-shaped pattern between solar radiation and cases meaning that the solar radiation would be able to reduce the number of infected people only for high levels of radiation. Regarding the fatality rate regressions (SI Appendix, Table 11), solar radiation as well as temperatures exhibit significant threshold effects. These results reveal that the solar – nonlinear – radiation effect has been probably underestimated in previous literature. These nonlinearities might also be explained by some others factors like the mobility of households and workers.

### Indirect climatic factors effects and mobility

We find (Tables 2-3 and SI Appendix, Tables S12-S15) that the mobility *driving* variable has a direct negative influence on Covid-19 outcomes in cases regressions but a positive or null influence in the fatality rate regressions. Indeed, DFE estimates show that the mobility variable has a significant and positive correlation with the Covid-19 fatality rate (Table 3). As a consequence, an increasing mobility would increase the fatality rate of the virus, confirming that lockdown policies have been probably relevant to try to reduce the severity of the pandemic. Again, this result can also be explained by the fact that the fatality rate captures a different time horizon in comparison with the infected cases ratio. Additionally, most countries in our sample have implemented a lockdown policy: in the short-run, the effect of this policy can be influenced by reverse causal endogeneity issues: people were asked to reduce their mobility (driving or walking, for instance) due to the strong diffusion of the virus. A increased number of cases has thus been associated with a reduced mobility. We control this potential endogeneity issue and our tests reveal that when the number of lags is increased (SI Appendix, Tables S16-19), the sign of the mobility variable turns to be positive also in the infected cases regression. In other words, increasing individual mobility is a factor of virus spread: when more people are more mobile, the social distancing is likely to be reduced and the transmission rate to increase.

**Table 2.**
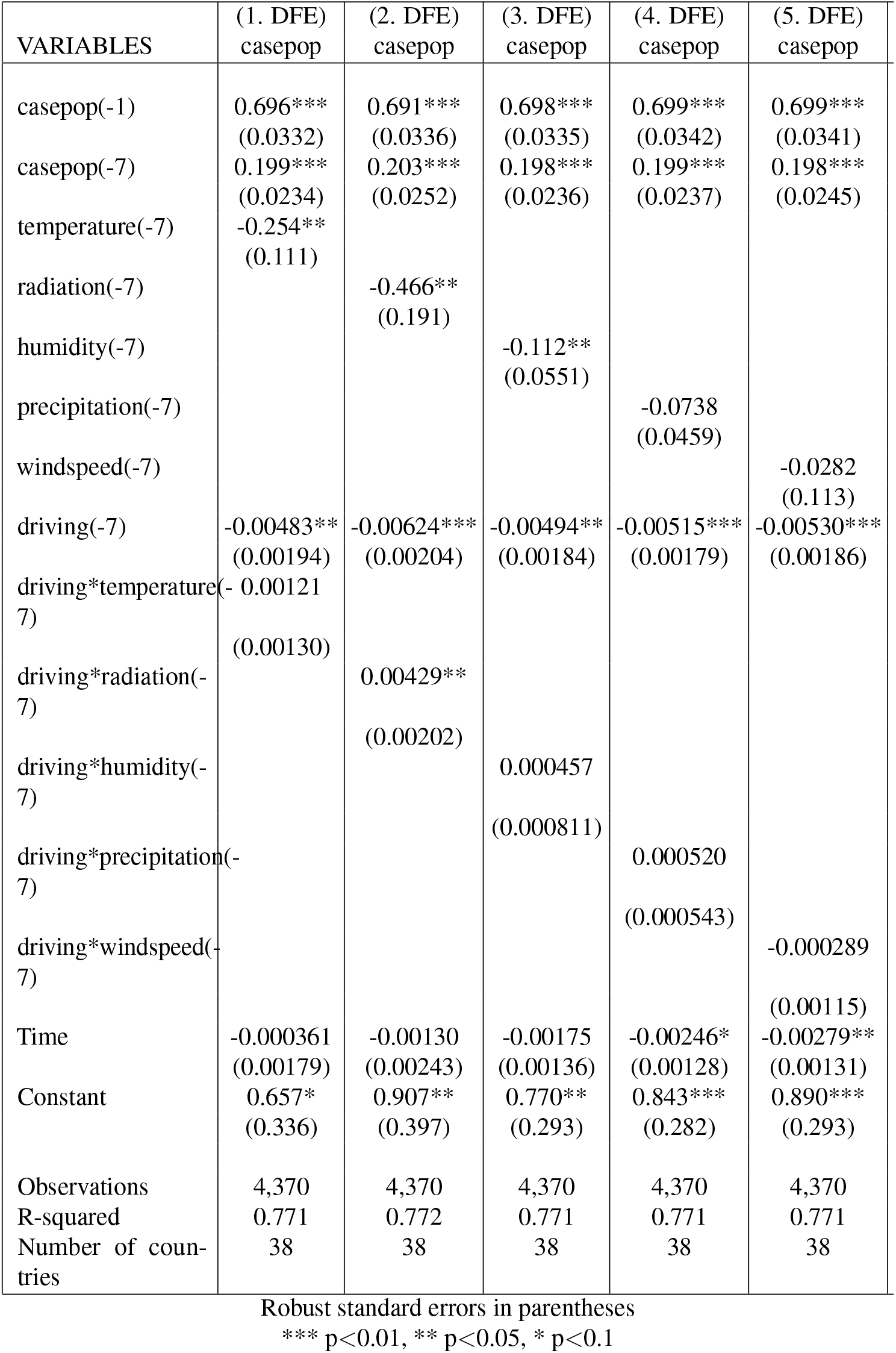
Indirect climatic effects on Covid-19 cases via social mobility

**Table 3.** Indirect climatic effects on Covid-19 fatalities via social mobility: DFE

| VARIABLES | (1. DFE)<br>deathpop | (2. DFE)<br>deathpop | (3. DFE)<br>deathpop | (4. DFE)<br>deathpop |
| --- | --- | --- | --- | --- |
| deathpop(-1) | 0.623***<br>(0.0774) | 0.621***<br>(0.0770) | 0.625***<br>(0.0775) | 0.626***<br>(0.0764) |
| deathpop(-14) | 0.106**<br>(0.0420) | 0.109**<br>(0.0412) | 0.105**<br>(0.0416) | 0.108**<br>(0.0413) |
| casepop(-14) | 0.0109**<br>(0.00470) | 0.0105**<br>(0.00461) | 0.0111**<br>(0.00476) | 0.0112**<br>(0.00481) |
| temperature(-28) | -0.0326**<br>(0.0148) |  |  |  |
| radiation(-28) |  | -0.0456***<br>(0.0148) |  |  |
| humidity(-28) |  |  | -0.0155<br>(0.0143) |  |
| precipitation(-28) |  |  |  | 0.00661<br>(0.00629) |
| driving(-28) | 0.000659**<br>(0.000270) | 0.000583**<br>(0.000255) | 0.000648**<br>(0.000276) | 0.000646**<br>(0.000273) |
| driving*temperature(-28) | 0.000255*<br>(0.000129) |  |  |  |
| driving*radiation(-28) |  | 0.000440**<br>(0.000166) |  |  |
| driving*humidity(-28) |  |  | 0.000123<br>(0.000108) |  |
| driving*precipitation(-28) |  |  |  | -3.29e-05<br>(6.90e-05) |
| Time | 0.000716**<br>(0.000272) | 0.000718***<br>(0.000227) | 0.000603**<br>(0.000256) | 0.000580**<br>(0.000230) |
| Constant | -0.0985**<br>(0.0473) | -0.0866**<br>(0.0413) | -0.0881*<br>(0.0471) | -0.0865*<br>(0.0447) |
| Observations | 3,914 | 3,914 | 3,914 | 3,914 |
| R-squared | 0.563 | 0.564 | 0.563 | 0.563 |
| Number of countries | 38 | 38 | 38 | 38 |

Robust standard errors in parentheses
For space reasons, insignificant windspeed regression is not reported
\*\*\* p<0.01, \*\* p<0.05, \* p<0.1

When we look at the infected cases with a short-run 7 days delayed effect, we find that the interaction term *driving* * *radiation* is significantly positive but other interaction terms are insignificant. In others words, during sunshine periods, the mobility effect on the number of infected cases is strengthened by this climatic factor within a 7 days action period. Nonetheless, these results are more significant when considering the fatality rate as a dependant variable and a 28 days lagged period for the effect of climate and mobility variables. This could be explained by the higher relevance of the fatality rate variable in comparison to the infected cases one to capture the true dynamics of the pandemics. We derive significant results mostly for solar radiation although temperatures should also be considered.

Climatic factors – solar radiation here – have always a negative direct coefficient since solar radiations are expected to physically reduce the resistance of the virus. The interaction term between solar radiation and mobility is positive and shows that both variables strengthen each other. Since climate is *per se* purely exogenous, climate tend to strengthen the effect of the mobility on the virus spread. The net effect from climate on Covid-19 outbreak will thus result from its direct negative effect and from its indirect positive effect resulting from the interaction between mobility and climate variables. We show (Table 2 and SI Appendix, Tables S12-S13) that there is a direct negative effect of the mobility on the infected cases ratio – we probably capture the lockdown period – but there is an indirect effect of solar radiation on *casepop* through the interaction between solar radiation and mobility: a highest sunshine duration is associated with a reduced number of infected cases. A combination of increasing mobility and solar radiation will lead to an increasing number of infected cases. Our results (Table 3 and SIAppendix, Tables S14-S15) reveal the existence of a direct positive effect of the mobility variable on the fatality rate but a direct combined negative effect of temperatures and solar radiation on the fatality rate through the mobility. As a consequence, high levels of temperatures and sunshine are likely to reduce the fatality rate by physical channels. However, this potential reduction is however partially compensated by a significant positive effect stemming from the interaction between climatic variables – temperatures and solar radiation – and the mobility variable, on Covid-19 fatality rate.

## Discussion

Understanding links between climate and Covid-19 is crucial. If climatic factors have a physical impact on the virus, it would be helpful and a supplementary weapon to stop the virus in complement to sanitary policies implemented by the authorities.

Our results reveal that the choice of delays and time perspective of the effects of climatic factors on the virus as well as the choice of Covid-19 outcomes are crucial and can explained the disparate findings reported in the previous literature. Concerning direct effects of climate on Covid-19, our work highlights that temperatures and, more interestingly, solar radiation – that has been undervalued in previous studies – are weak significant climatic drivers on Covid-19 outbreak. Climatic factors – solar radiation here – has always a negative direct coefficient since UV are expected to physically reduce the resistance of the virus. Others climatic factors seem less prone to impact Covid-19 outcomes.

Our estimates point that direct climate effects are not completely clear due to several econometric issues and, above all, that indirect effects through human behaviors should be investigated simultaneously. In addition, interrelationships between some climatic variables and mobility are significantly positive. Since climate is *per se* purely exogenous, climate tend to strengthen the effect of mobility on the Covid-19 virus spread. The net effect from climate on Covid-19 outbreak will thus result from some potential direct negative effects of climatic variables and from indirect effects through changes in human behaviors in relation with climatic conditions. Our results show that direct negative effects from climatic (temperatures and solar radiation) factors on Covid-19 outcomes – when they are significant – are partly compensated by positive indirect effects through human mobility.

This does not mean that climate has a neglected role in Covid-19 epidemics. This simply means that climate has not a major direct effect, though significant, *via* physical channels. In addition, climate can influence other crucial determinants such as social distancing, wearing mask through mobility and human behaviors. Indeed, meteorological factors are drivers of the mobility of lot of people. In one side, high temperatures and sunshine lead people to move away and do not stay in clustering places that could reduce the transmission rate by introducing more distance between people since it is now well known that the virus is essentially transmitted through droplets generated *via* coughing and sneezing and in a less extent through the air, when tiny particles, or aerosols, hang around. A minimum one meter distance is requested but a 4 meter distance has also been suggested very recently (36). Climatic factors can thus reduce the spread of the virus through this indirect channel. On the other side, when climate conditions are favorable and people are spending time outdoors, they can go in the parks, in meetings, in terraces; this can lead sometimes to careless behaviors when people reduce social distancing or do not wear masks when it is necessary.

*What implications about policy?* Finally, a hot summer could only slightly enhance the effect of stringent social distancing. Thus, high temperatures are not substitute to suitable hygienic and social distancing measures. As a consequence, suitable hygienic and public policies would be consist in mandating the mask wearing and reduce temporarily the individual mobility.

*What implications of this work for the future?* This work could be extended by introducing air quality and pollution factors as potential supplementary drivers in our model since it is likely that polluting activities can increase the virus intensity. More generally, investigating relationships between climate, meteorological factors and viruses is important to understand how climate change might influence epidemics dynamics and outbreaks in the coming years. Recently, a UNO report (37) untitled "Preventing the next pandemic" conclude that zoonetic diseases are likely to increase in the near future. Climate change and unsustainable human activities are drivers of the increasing frequency of pathogenic microorganisms jumping from animals to people. Beyond the link between climate and Covid-19 pandemic, it is necessary to better investigate the links between climate and human behaviors and the link between climate and moving borders between humans and animals that are in the epicenter of the disease transmission.

## Methods

### Data and variables

We conducted an original empirical work based on a 42 countries dataset between the first of January and the 28th of May 2020. We consider 5 different meteorological factors and do not focus only on temperatures as in many previous studies. We obtain a panel with 42 countries and more than 4600 observations. We obtained (1) the number of confirmed COVID-19 cases and deaths for the countries in our sample from multiples sources through the DELVE initiative between 1st January 2020 and May, 27th 2020, (2) estimated population in 2019 from the World Bank’s World Development Indicators database, and (3) daily meteorological conditions in the selected countries for 6 months. Covid-19 data have been aggregated by (28), DELVE Global COVID-19 Dataset and are available at http://rs-delve.github.io/data_/global-dataset.html.

We used both Covid-19 infected cases and mortality data per capita. One reason behind is that cases counts can be biased (29). In some countries, during the pandemic waves, testing has been stopped due to lack of time concerns. Another reason to consider both variables is they do not capture the same Covid-19 outcomes. Infected people are generally officially considered as infected (by testing) only few days (between 7 and 14 days) after the contamination day although the median incubation is estimated to be 5 days (30). Infected cases counts thus captures a short-run effect of potential climatic or social distancing variables on the Covid-19 pandemic. However, the delay is longer for deaths counts considering the time people can develop the disease and stay in hospitals. As a consequence, the fatality rate captures a long-run face of the transmission from climatic and other determinants to Covid-19 outcome.

Most selected OECD countries in our sample have encountered the first wave of the pandemic quite in the same time (SI Appendix, Table 32). We take into account the huge numbers of zero values for the infected cases and the fatality rate (especially for this latter) in the beginning of the sample range by choosing a restrictive sample as a benchmark: February 1st has been assumed as a starting time for the pandemic (infected cases) for all countries whereas February 15th has been chosen for fatality counts. We have considered epidemiological and also statistical (lags) arguments when we selected these particular dates. Robustness checks (SI Appendix) show that the heterogeneity of the starting dates is not a significant issue.

Concerning the climatic factors, even though there exist significant differences in meteorological conditions between our set of countries, they are all located in a relative homogeneous North Hemisphere. We thus have all conditions to work with a suitable panel: unobserved heterogeneity across countries is included inside a relative homogeneity panel (see the “to pool or not pool debate”, for instance in (31)).

Meteorological conditions are summarized (SI Appendix, Table S2) as: mean daily temperatures (in Celcius degrees), total precipitation (mm / hr), mean wind speed (mph), solar radiation (Watts per square meters) and mean relative humidity (Kilograms of water vapour per kilogram of air). For each country, all climatic observations have been weighted by the population. Note that the five meteorological factors have been standardized for comparisons purposes and for coefficients scale homogeneity.

Finally, we use Apple mobility reports data. Two main variables are used to proxy the people mobility: *mobility_worplaces* et *mobility_residential*. These data are indexes (100 basis) and are computed as the % change in routing requests since 13th January 2020. Note (see more details in the data section) that the mobility index (from Apple) takes low values when mobility is low and people prefer stay indoors and high values when the mobility (driving or walking) is stronger. A January basis (before the pandemic) has been considered as a statistical benchmark scale.

### Theoretical assumptions

Climate patterns are tested by investigating the significance of coefficients associated to five climatic factors: temperatures, humidity index, precipitations levels, wind and solar radiation. Expected results and assumptions are summarized in the Table 4: — denotes negative effect ie a decreasing number of cases or deaths and + a positive effect ie an increasing number of cases or fatalities. We also use the ? symbol denoting that there exist some uncertainty regarding the absence of significant investigation about this question or mixed results. Temperatures are expected to directly impact the virus negatively but indirect effects by increasing incentives for people to go outdoors may be negative at low levels or positive at higher levels. Precipitations have no expected direct effect but indirect impact are expected; indeed, too much rains could generate incentives to stay indoors and increase the transmission of the virus. Humidity has been identified as a direct negative driver in some lab experiments but its indirect effect through human behavior is difficult to evaluate. The same observation stands for Wind. Solar radiation is likely to reduce the durability of the virus and its indirect effects are expected to be relatively similar to those associated with temperatures. Note in a general manner that mobility is not related to weather in an homogeneous manner. Activities that are done for leisure purpose are conditional to climatic conditions whereas mobility for work purpose for instance is an incompressible task that is *de facto* not sensitive to climatic conditions.

**Table 4.** Expected effects of climatic factors on Covid-19 outcomes

| Variables/Effect | Direct | Indirect |
| --- | --- | --- |
| Temperatures | - | - or + |
| Precipitations | null or ? | + |
| Humidity | - | ? |
| Wind | - or ? | ? |
| Solar | - | - or + |

### Dynamic econometric panel model

To test our theoretical assumptions, we use a dynamic panel econometric model as follows:

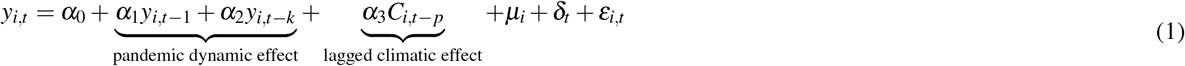

Where the subscripts *i* and *t* represent country index and periods (days) respectively. The dependent variable, *y_i_,_t_*, can be the number of infected people (casespop) or deaths (deathpop) per capita (considering the population size) at time t. *C_i_,_t−p_* is a vector of variables depicting the effects of meteorological conditions in day *t _−_ p*. Country-specific fixed effect, *µ_i_*, are included to control for time-invariant omitted-variable bias and *ε_i_,_t_* is the error term. *δ_t_* is a deterministic time trend that controls the deterministic dynamics of the epidemics over the studied period and captures some unobserved information about the pandemic common to all countries. In addition, lagged terms *y_i_,_t-k_* capture the stochastic part of the pandemic dynamics. We assume *k* equal to 7 or 14 lags/days in our baselines specifications considering incubation and confirmation periods presented in the Covid-19 literature. Moreover, for logical reasons, since the climatic factors do not immediately impact the Covid-19 spread, the climatic variables are also included in our model with a lag of order *p*. Indeed, there are delays between the time of potential infection corresponding to certain climatic events and the time of official counting of a potential infected people (or fatality). Therefore, when dealing with p, 7 or 14 days are considered when *case pop* is used as endogeneous variable considering the short period between transmission and infection. A benchmark 28 lags (about one month) delay is considered when dealing with *deathpop* because of the more important lag lenght assumed between the infection and the deaths related to the Covid-19 virus. More lags have been also considered in robustness checks to take into account the dynamic persistence of the pandemic (SI Appendix). *casepop* and *deathpop* respectively give a short-run and medium/long-run time perspective of the dramatic outcomes of the pandemic. Note that when *y_i,t_* is the fatality rate, we also add the ratio of infected cases per capita in our benchmark specification in order to account for the fact that the level of the pandemic can impact the fatality rate. The reason behind is to control for a level effect and a kind of saturation effect of the health system (too many infected people to manage is likely to finally increase the fatality rate).

This baseline specification is extended to take into account different potential nonlinearities: quadratic terms for climatic variables (to capture thresholds effects), interacted terms between climatic variables are also added in the model due to high levels of multicollinearity. More importantly, our extended specification incorporates mobility indexes to investigate potential indirect effects of the meteorological factors via the impact of climate on human behaviors. Indeed, as stressed by (9), a substantial omitted variable bias is likely to result if mobility is not considered. (10) estimate the impact of mobility on the transmission rate of the virus using a Bayesian framework that captures the impact that non-pharmaceutical interventions and other behaviour changes have on the rate of transmission of SARS-CoV-2. Assuming a 14 lags delay, (32) also estimated the impact of mobility on the growth of infected people and found that a 10% decrease in mobility is associated with a 14.6% decrease in the average daily growth rate of cases.

Thus, equation (1) becomes equation (2) with *M* a mobility index:

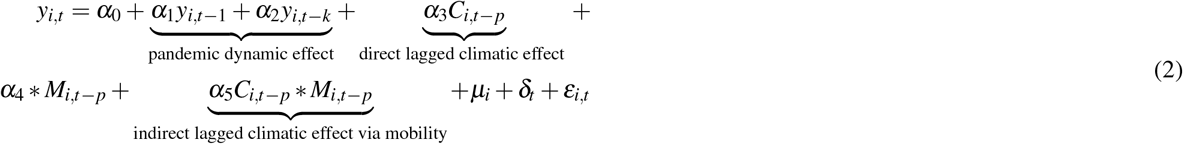

### Identification issues

Equations (1) and (2) can be estimated by the mean group (MG) and the dynamic fixed effect (DFE) estimators introduced by (33) and (34). Both estimators are relevant for macro panels such as the one used in this paper: *T* is equal to 148 and is thus largely superior to *N =* 42 (see SI Appendix). We also used Syst-GMM estimates and many robustness regressions and tests to validate the consistency of our results (SI Appendix, Section 6). The Mean Group (MG) estimator consists in estimating each regression separately for each panel member *i* (country here) with a minimum of restrictions. All estimated coefficients are heterogeneous and are subsequently averaged across countries *via* a simple unweighted average (33). An intercept is included to capture country fixed effects as well as a linear trend. In the dynamic fixed effects (DFE), the slopes are homogeneous but the intercepts are allowed to vary across countries (34).

Although we apply appropriate macro-panel estimators to our data, several issues can nonetheless emerge. First, using dynamic models, we are vulnerable to the so-called Nickel bias. Here, this bias is relatively negligible, notably considering the important time length of our series. Second, panel regressions may be exposed to an omitted variables bias. It would be possible to include control variables such as control measures (e.g. testing, mask wearing, travel controls) or structural determinants (e.g. population density and demographics such as the population over 65, tourists flows, GDP per capita, and measures of health infrastructures). Considering the so-called problem of controls in the so-called “climate macroeconomy literature”, our set of explanatory variables is assumed to be restricted to climatic variables in order to avoid an over-controlling problem (35). In addition, considering data availability and the fact they are time-invariant variables, we capture these unobservables via the lagged term *y_i_,_t−_*_1_ and above all with country fixed effects. Another identification issue is related to the potential reverse causality bias related to our Covid-19 variables: news about contemporaneous dynamics of the Covid-19 outbreaks and counts can change the human behavior in real time and the social distancing. This is why lags of dependent variables must be added in our model. Finally, persistence and multicollinearity are other usual issues in panel studies. We have controlled for both by computing autocorrelations LM (Lagrange Multiplier) tests and VIF/Tolerance ratios after each estimated regression. In SI Appendix, we also consider endogeneity issues about the mobility index variable and other tests related to the specification of our econometric model, the choice of an alternative estimator, and several changes in the sample composition.

## Data Availability

Data are available upon request and on a public website.

## Acknowledgements (not compulsory)

We are grateful to L. Muehlenbachs, Co-Editor of the Journal of Environmental Economics and Management, for encouragements to submit our paper in a *Science* journal and to May Eichenbaum, editor of PNAS.

## Author contributions statement

Conceptualization, Data curation, Formal analysis, Investigation, Methodology, Project administration, Software, Supervision, Validation, Visualization, Writing – original draft an Writing – review editing by O. Damette, Formal analysis, Investigation, Methodology, Software, Writing – review editing by C. Mathonnat, Conceptualization, Data curation, Investigation by S. Goutte.

No conflict of interest.

## Corresponding author

The correspondence should be addressed to olivier.dametteuniv-lorraine.fr; The corresponding author is responsible for submitting a [competing interests statement](http://www.nature.com/srep/policies/index.html#competing) on behalf of all authors of the paper.

## Additional information

**Accession codes:** SI dataset in.dat format and Stata codes (do files) are attached with the submitted manuscript and available at: https://sites.google.com/site/olivierdamette/research?authuser=0

## Notes

### Competing Interest Statement

The authors have declared no competing interest.

### Funding Statement

No Funding.

### Author Declarations

Not relevant here.

